# Subject-Disjoint, Imbalance-Aware Gameplay–Rest Classification in Pediatric Mobile EEG Using Enriched Topological Features

**DOI:** 10.1101/2025.11.05.25339645

**Authors:** Waleed Bin Owais, Herbert F. Jelinek, Ibrahim (Abe) M. Elfadel

## Abstract

**Objective:** Classifying cognitive state from pediatric mobile electroencephalography (EEG) recorded during naturalistic behavior is challenging under severe class imbalance, subject variability, motion artifacts, and low channel count. To avoid inflated performance estimates, this study enforces subject-disjoint evaluation and imbalance-aware metrics to distinguish Minecraft gameplay from eyes-open rest.

**Methods:** We propose Enriched Topological Features (ETF), which combine persistence landscapes with temporal entropy aggregation within Takens phase-space embeddings derived from persistent homology. ETF was evaluated on four-channel Muse EEG from 43 children using subject-disjoint five-fold GroupKFold cross-validation. Class imbalance was handled exclusively within training folds using synthetic minority oversampling or within-subject random undersampling, while test folds preserved natural class proportions. Sensitivity of topological parameterization was assessed across persistence landscape resolution and truncation depth using linear mixed-effects modeling.

**Results:** Under oversampling-based training, ETF achieved test balanced accuracy up to 65.39% and macro-F1 up to 64.48%, with minority-class (rest) recall reaching 49.51% using only TP9/TP10 electrodes. Relative to persistence-landscape-only representations, ETF improved macro-F1 by approximately 6–10 percentage points under subject-disjoint, imbalance-aware evaluation. Subject-level bootstrap analysis confirmed that performance trends were not driven by unequal epoch counts per subject. End-to-end processing required 676 ms per 4.5 s EEG epoch on CPU, supporting near–real-time feasibility.

**Conclusion:** ETF enables imbalance-aware gameplay–rest discrimination from low-density pediatric mobile EEG under subject-disjoint evaluation.

**Significance:** These results demonstrate that enriched topological representations remain informative under realistic motion and hardware constraints, supporting coarse engagement–rest state monitoring in pediatric mobile neuroengineering settings.

## 1. Introduction

Understanding the neural basis of children’s behavior in real-world settings remains a significant challenge in developmental neuroscience. Young children acquire cognitive, language, and social skills through dynamic, natural interactions, yet most neurophysiological studies still occur under solitary and highly controlled laboratory conditions [1]. Classifying pediatric electroencephalography (EEG) during naturalistic activities such as free play or gameplay with unconstrained movement offers more ecologically valid insights into brain function that are directly relevant to children’s everyday experiences [1].

Advances in wearable EEG technology have made it possible to record brain activity outside of traditional laboratory environments [2]. This includes contexts where conventional participation is difficult, such as children with autism who may find lab visits stressful or families in remote communities. These developments reflect a growing effort to bridge the gap between laboratory-based protocols and real-world pediatric neuroscience. Conventional EEG paradigms, however, often fall short of this goal. Most developmental EEG studies restrict child movement and minimize environmental complexity, typically instructing children to remain seated and view repetitive screen-based stimuli [1]. Adding to the challenge, children, especially infants and toddlers, are naturally active. Studies show that even during “calm” screen-based tasks, children move during the majority of recording time [3]. Such movement introduces artifacts that are difficult to suppress without either discarding large portions of data or removing meaningful neural signals [3]. These limitations highlight the need for alternative methods capable of tolerating motion while preserving signal interpretability.

To address these challenges, Mobile Brain–Body Imaging (MoBI) and mobile EEG systems have emerged as effective tools for studying brain activity during real-world behavior [4]. MoBI combines EEG with motion capture or inertial sensors, enabling participants to move freely while cognitive processes are recorded in context [5]. However, ecologically valid pediatric EEG datasets acquired from freely behaving children in dynamic, non-clinical environments remain rare, limiting systematic evaluation of classification methods under realistic motion and noise conditions.

This study enforces three evaluation constraints that are commonly not explicitly enforced in prior work: (i) subject-disjoint evaluation, (ii) preservation of natural rest–task imbalance in the test data, and (iii) imbalance-aware metrics as primary performance outcomes. EEG recorded from children engaged in naturalistic video game play is analyzed to assess whether task-related neural structure can be recovered under high-movement conditions using low-density mobile recordings. In doing so, we quantify what gameplay–rest discrimination performance remains once two common sources of inflation are removed, that is subject leakage and accuracy-dominated reporting under severe rest– task imbalance. Under these deployment-relevant constraints, overall accuracy is no longer an appropriate benchmark; instead, class-balanced discrimination and minority-class (rest) sensitivity provide a more meaningful assessment of model performance. This framing establishes a deployment-relevant baseline for pediatric mobile EEG analysis, rather than an optimized laboratory benchmark.

### Contributions

This work makes four primary contributions. First, we define a deployment-relevant evaluation protocol for pediatric mobile EEG that enforces subject-disjoint testing, preserves natural rest–task imbalance in held-out folds, and treats imbalance-aware metrics as primary outcomes. Second, we introduce **Enriched Topological Features (ETF)**, which enhance persistence landscape representations by incorporating persistence entropy and temporal aggregation. Unlike prior methods such as NETDA, ETF explicitly couples persistence landscapes with diagram-level entropy and temporal aggregation, stabilizing minority-class decision boundaries under severe imbalance. This makes ETF a deployable, imbalance-aware protocol specifically designed for pediatric MoBI with low-density, high-motion EEG recordings. Third, we conduct a pre-specified sensitivity analysis of key topological parameters (persistence landscape resolution and truncation depth) using linear mixed-effects modeling. Finally, we quantify interpretability and deployment feasibility by reporting SHAP-based feature attribution and end-to-end CPU runtime under both two-channel and four-channel Muse configurations.

## II. Related Work

Conventional EEG analysis approaches for cognitive state classification often rely on spectral power measures, frequency decomposition, and functional connectivity metrics. While well-established, these methods can degrade under nonstationarity and nonlinear dynamics, common in naturalistic and task-evoked EEG recordings [6]. These limitations are especially pronounced in pediatric EEG, where motion artifacts, high inter-subject variability, and limited durations of artifact-free data further degrade signal reliability and model generalization.

Neural network-based models, including convolutional architectures, have been applied to EEG to directly learn task-related representations. However, their deployment in MoBI settings is constrained by practical challenges such as low-channel EEG systems, small cohort sizes, and substantial recording noise [7]. Studies aiming to distinguish resting from task states under similar mobile and pediatric conditions have reported only moderate performance, reflecting the difficulty of extracting stable neural signatures under these constraints [7].

EEG microstate analysis has been proposed as an intermediate-scale representation of brain dynamics, modeling EEG as sequences of quasi-stable spatial patterns. Microstate temporal statistics and transition structures have been investigated as biomarkers across various neurological and psychiatric conditions, including schizophrenia, tinnitus, trigeminal neuralgia, and major depressive disorder [8]–[11]. While these approaches capture temporal organization beyond static spectral summaries, they discretize neural activity into a small set of states and primarily characterize dynamics through state occupancy and transitions. This limits their ability to represent continuous, multiscale temporal structure, particularly in noisy, low-density mobile EEG recordings. Additionally, reliable microstate estimation generally requires higher spatial sampling than is available in low-density mobile EEG systems, limiting applicability in this context.

Beyond discrete-state representations, nonlinear time-series analysis methods aim to characterize EEG dynamics through measures such as entropy, fractal scaling, and phase-space reconstruction. These methods offer insights into higher-order temporal organization but often exhibit sensitivity to parameter selection and limited robustness across subjects and recording conditions [12].

Recent advances in topological signal processing (TSP) have introduced noise-tolerant frameworks for characterizing the geometric structure of neural dynamics [13]. By applying persistent homology to phase-space embeddings of EEG signals, TSP captures invariant topological features that reflect global dynamical organization rather than localized signal properties. This approach has demonstrated utility in applications such as neurodegenerative disease and cognitive-state classification in complex, naturalistic scenarios [14].

A notable application of this paradigm is the Nonlinear EEG Topological Dynamics Analysis (NETDA) framework [15], which applies topological data analysis to pediatric EEG recorded during real-world gameplay. NETDA achieved high accuracy under accuracy-centric evaluation by extracting persistence-based features from reconstructed phase spaces. Its effectiveness stems from the noise tolerance of topological representations and their suitability for low-channel EEG. However, NETDA is sensitive to embedding parameter choices, provides limited interpretability of extracted features, and does not address class imbalance or deployment constraints explicitly. These limitations highlight the need for further refinement of topological approaches to enable robust and interpretable cognitive state classification in pediatric mobile EEG.

Despite these advances, several gaps remain in applying topological methods to pediatric mobile EEG. Prior work has often emphasized accuracy-focused evaluation in the presence of severe class imbalance, frequently without reporting balanced accuracy or minority-class sensitivity. Additionally, existing frameworks offer limited analysis of parameter robustness, minimal insight into the interpretability of extracted topological features, and infrequent reporting of computational feasibility for online or near–real-time use.

These gaps motivate the development of a topological representation and evaluation protocol designed for subject-disjoint testing and severe rest–task imbalance.

## III. Topological Foundations of Feature Extraction

### A. Vietoris–Rips Complexes and Simplicial Filtration

Topological Data Analysis (TDA) offers a formalism for capturing the intrinsic structure of data through topological invariants. Central to this framework is the notion of a *simplicial complex*, a combinatorial structure that generalizes graphs to higher dimensions. A *k*-simplex consists of *k* + 1 affinely independent points; examples include 0-simplices (vertices), 1-simplices (edges), 2-simplices (triangles), and so on [16].

For a finite point cloud *X* ⊂ ℝ^*d*^, the Vietoris–Rips complex ℛ (*X*, ϵ) at scale ϵ > 0 is defined as the set of simplices σ ⊂ *X* for which all pairwise distances between points in σ are at most ϵ:

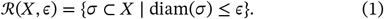

As ϵ varies, these complexes form a filtration that encodes the data’s shape across multiple spatial resolutions.

### B. Persistent Homology and Multiscale Topology

The evolution of topological features across a filtration is quantified using persistent homology. Each topological structure, such as a connected component or loop, is tracked by its birth and death scales, forming a homology class ℍ_*k*_, where ℍ_0_ corresponds to connected components and ℍ_1_ denotes one-dimensional cycles [17].

Each feature is represented as a birth–death pair 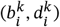, with persistence (or lifetime) defined as

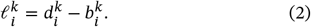

The collection of all such pairs forms a persistence diagram, which summarizes the multiscale lifespans of topological features. In this work, analysis is restricted to the first two homology dimensions, ℍ _0_ and ℍ _1_, which capture coarse connectivity and loop-like structure in reconstructed EEG phase-space trajectories.

### C. Topological Descriptors: Landscapes and Entropy

While persistence diagrams are extensive in information, they are not directly amenable to vector-based analysis. To overcome this, persistence landscapes transform diagrams into functions in a Hilbert space, enabling consistent and differentiable representations [18]. These are constructed by projecting birth– death intervals onto piecewise-linear functions, retaining the most significant layers for further analysis.

In addition, persistence entropy [19] serves as a compact summary of diagram complexity. For a persistence diagram *D* = {(*b*_*i*_, *d*_*i*_)}, the persistence entropy is given by:

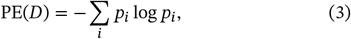

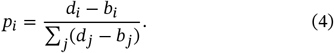

This measure reflects the spread and uniformity of topological lifespans within a given homology dimension and provides a scalar descriptor of structural regularity. Persistence landscapes preserve multiscale geometric structure, while persistence entropy provides a compact summary of diagram complexity; together, they offer complementary representations of topological organization.

## IV. Dataset Description

This study uses the publicly available Mobile Brain–Body Imaging (MoBI) dataset collected at the Children’s Museum of Houston [20]. EEG recordings were available from 171 children (ages 6–16 years; mean age 8.7 ± 2.2) in the publicly released dataset. Each participant completed an eyes-open resting period (approximately 1 minute) followed by up to 20 minutes of free-play Minecraft gameplay in a real-world environment.

EEG signals were recorded using a 4-channel Muse headband (TP9, TP10, AF7, AF8) at a sampling rate of 220 Hz with an online 60 Hz notch filter. Concurrently, 3-axis head motion was captured via onboard accelerometers at 50 Hz. The dataset includes raw EEG signals, inertial sensor recordings, and event markers intended to enable segmentation of resting and gameplay periods.

All EEG and accelerometer recordings were obtained directly from the publicly released `.mat` files provided by the dataset authors. No proprietary Muse preprocessing software (e.g., Muse SDK or MuseDirect) was used. All preprocessing, epoch segmentation, quality control, and subject inclusion criteria were implemented using a custom analysis pipeline, as described later.

### A. Subject Attrition and Inclusion Criteria

A total of 171 EEG recording files were available in the public MoBI dataset. Of these, 88 recordings contained explicit baseline and gameplay segmentation markers within the EEG structure. The remaining 83 recordings were excluded at this stage due to missing segmentation metadata, which precluded reliable separation of resting and gameplay intervals.

Among the 88 recordings with segmentation markers, 7 were excluded due to invalid or malformed marker indices (e.g., out-of-range or improperly ordered baseline or task boundaries), yielding 81 subjects with valid baseline and gameplay segments. All 81 of these subjects contained sufficient data to form at least one 4.5 s epoch in both conditions prior to quality control.

Following epoch-level quality control, described in Section V-A, which included amplitude-based rejection and spectral contamination screening, 46 subjects retained at least one quality-controlled resting epoch and 56 subjects retained at least one quality-controlled gameplay epoch. A total of 43 subjects retained at least one quality-controlled epoch in both conditions and were included in the final paired analysis. Subject exclusion was driven exclusively by missing segmentation metadata or failure to retain any quality-controlled epochs in one or both conditions, and not by classification performance or outcome-dependent criteria. A summary of subject attrition across processing stages is provided in Supplementary Table S3.

### B. Ethical Approval

This work constitutes a secondary analysis of a publicly available, fully de-identified pediatric EEG dataset collected as part of the Mobile Brain–Body Imaging (MoBI) project at the Children’s Museum of Houston. The original data collection protocol was approved by the Institutional Review Board (IRB) at the University of Houston. Written informed consent was obtained from parents or legal guardians, and informed assent was obtained from all participating children, as reported in the original dataset publications [20]. The dataset is openly available through IEEE DataPort (DOI: 10.21227/H23W88) and contains no personally identifiable information. No new data were collected, and no interaction with human participants occurred in the present study.

## V. Methodology: Feature Extraction and Classification with Enriched Topological Features (ETF)

The Enriched Topological Features (ETF) framework is designed to support classification of cognitive brain states from low-density pediatric EEG recorded in naturalistic environments. It combines topological signal processing with entropy-based temporal summarization to capture nonlinear patterns in brain activity while remaining stable under real-world noise and variability. To handle class imbalance and subject heterogeneity, ETF uses both random undersampling and synthetic oversampling methods. The pipeline is modular and interpretable, and was designed with computational efficiency in mind for mobile neuroimaging and time-sensitive applications. The method is summarized in Algorithm 1.

### A. Preprocessing and Signal Transformation

EEG data were acquired from four Muse headband channels (TP9, TP10, AF7, AF8) at a sampling rate of 220 Hz. For subjects meeting the inclusion criteria described in Section IV-A, resting and gameplay intervals defined by embedded event markers were extracted and processed using a standardized preprocessing pipeline prior to feature extraction.

Raw EEG signals were first high-pass filtered at 1 Hz using a zero-phase 4th-order Butterworth filter to remove DC offset and low-frequency drift. A zero-phase notch filter centered at 60 Hz was then applied to attenuate line noise contamination [21], followed by band-specific zero-phase FIR filtering corresponding to the analyzed frequency bands. Signals were demeaned per channel after filtering, and all subsequent phase-space reconstruction and topological analyses were performed on these band-limited, demeaned signals.

Continuous recordings were segmented into non-overlapping 4.5 s epochs. Each epoch was subjected to a multi-stage quality control procedure. Epochs exhibiting excessive amplitude fluctuations, defined as a standard deviation exceeding 75 *μ*V on any channel, were rejected. Epochs with abnormally low variance (standard deviation below 0.05 *μ*V), indicative of flatlined signals or sensor detachment, were also excluded. To mitigate electromyographic (EMG) contamination, power spectral density (PSD) was estimated using Welch’s method [22] on signals after high-pass (1 Hz) and notch (60 Hz) filtering but prior to band-specific FIR filtering. Epochs in which high-frequency power in the 35– 45 Hz range exceeded 25% of the total spectral power integrated over 1–45 Hz on any channel were discarded. The QC high-frequency band was chosen above the analyzed gamma range (30– 35 Hz) to detect high-frequency contamination without discarding physiologically relevant gamma-band content. All quality-control thresholds were fixed *a priori* and were not tuned to optimize classification performance.

Epoch-level quality control resulted in a final dataset of 3,788 retained epochs (369 resting, 3,419 gameplay) from subjects meeting the inclusion criteria described in Section IV-A, corresponding to an approximate class imbalance of 1:9.3. Cleaned epochs were retained in both a reduced two-channel configuration (TP9 and TP10) and the full four-channel configuration for downstream feature extraction and analysis.

### B. Phase-Space Reconstruction

Each band-limited EEG signal was transformed via phase-space reconstruction (PSR) using Takens’ embedding theorem [23] to recover the underlying state-space dynamics. To assess suitable phase-space embedding parameters, Average Mutual Information (AMI) [24] and False Nearest Neighbors (FNN) [25] analyses were used as descriptive tools to evaluate the stability of the time delay (*τ*) and embedding dimension (*m*) under the present preprocessing and epoching scheme. AMI estimated *τ* via the first local minimum of the mutual information curve, while FNN identified *m* as the smallest dimension for which the fraction of false neighbors fell below 10%.

Across subjects, channels, frequency bands, and behavioral segments, AMI- and FNN-derived estimates were highly concentrated, with a median delay of *τ* = 6 samples (interquartile range [6,6]) and a median embedding dimension of *m* = 4 (interquartile range [4,4]). These results indicate practical invariance of the embedding parameters under the present analysis settings. Accordingly, *τ* = 6 and *m* = 4 were fixed globally for all subsequent feature extraction and all cross-validation folds. *Train-only AMI/FNN estimates were invariant across folds*, and these parameters were not treated as tunable hyperparameters nor optimized with respect to classification performance.

This yielded embedded trajectories of the form

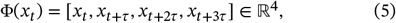

which preserve the geometric structure of the underlying temporal dynamics and serve as input to the topological analysis pipeline described in the following section. Representative AMI and FNN curves for a single subject are shown in Fig. 1. Subject- and channel-level AMI/FNN estimates, along with cohort-level summary statistics supporting this parameter choice, are reported in Supplementary Table S4.

**Fig. 1.**
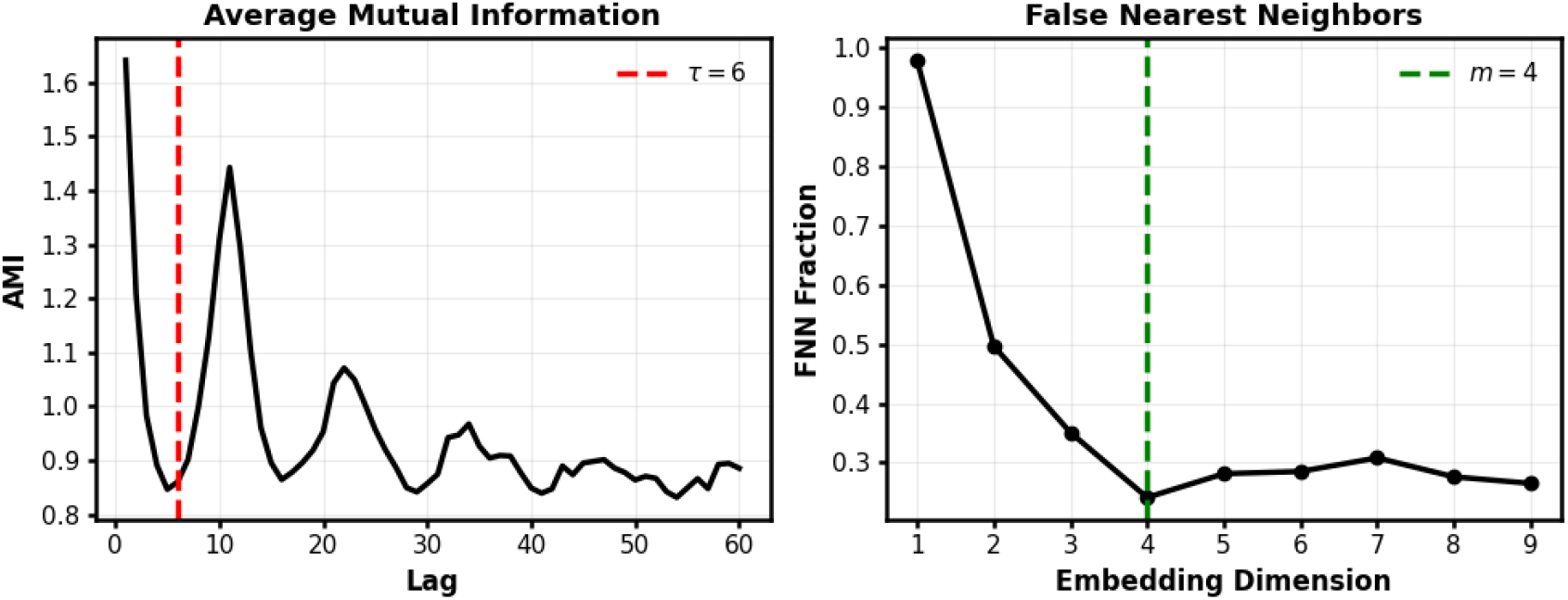
Estimation of phase-space embedding parameters using Average Mutual Information (AMI) and False Nearest Neighbors (FNN) analysis for a representative subject (Subject 27). (Left) AMI as a function of time delay, illustrating a pronounced local minimum in the range *τ* ≈ 5–6 samples. (Right) FNN proportion as a function of embedding dimension, exhibiting a sharp reduction at *m* = 4.

### C. Topological Feature Extraction

Topological data analysis (TDA) was employed to characterize the geometric structure of EEG dynamics. For each phase-space-reconstructed (PSR) trajectory, persistent homology was computed up to dimension one using Vietoris–Rips complexes, yielding persistence diagrams for connected components (ℍ_0_) and one-dimensional cycles (ℍ_1_).

Two complementary topological descriptors were extracted from each persistence diagram:

- **Persistence Landscapes (PL):** Birth–death intervals from ℍ _0_ and ℍ _1_ were transformed into persistence landscape functions, as described in Section III-C. Each landscape was sampled at PL = 30 uniformly spaced evaluation points to obtain fixed-length feature vectors.
- **Persistence Entropy (PE):** Each persistence diagram was summarized using persistence entropy, as defined in Section III-C (Eq. (3)–(4)), yielding a single scalar that quantifies the distribution of topological lifespans within each homology dimension.

#### Algorithm 1

*Feature extraction and classification with Enriched Topological Features (ETF)*

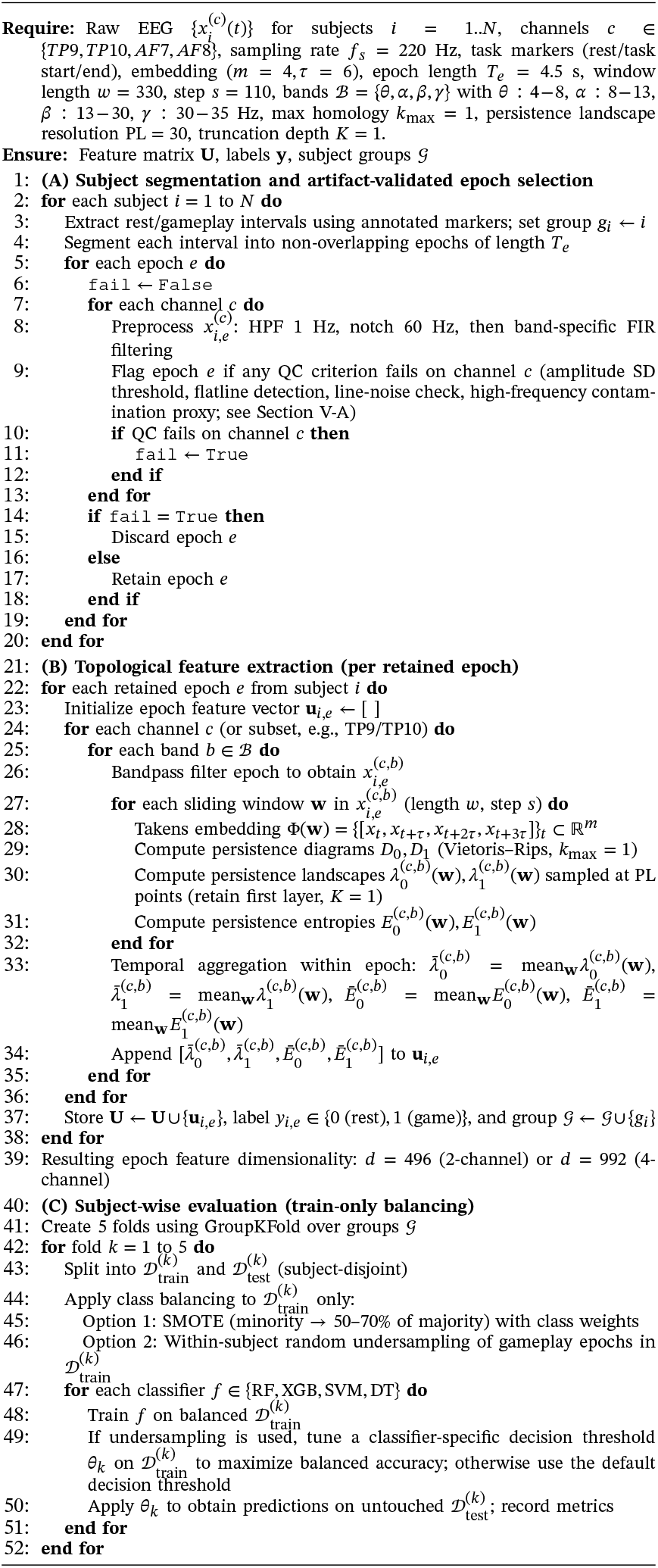

Each EEG epoch was represented by a fixed-length feature vector composed of persistence landscapes (30 values) and persistence entropy (1 value) computed across four frequency bands (theta, alpha, beta, gamma), two homology dimensions (ℍ_0_, ℍ_1_), and either two channels (TP9, TP10) or all four Muse channels (TP9, TP10, AF7, AF8). This resulted in feature dimensionalities of 496 and 992 for the two-channel and four-channel configurations, respectively. Features were computed within sliding windows and averaged over time to produce a single vector per epoch, capturing both geometric structure and topological complexity across multiple spatiotemporal scales.

To account for nonstationary signal behavior, a sliding-window approach was applied within each EEG epoch using a window length of 330 samples and a step size of 110 samples. Topological features were extracted per window and temporally averaged across all windows. Representative topological signatures derived from Subject 27 are shown in Fig. 2, illustrating differences in barcodes, persistence diagrams, and landscapes between gameplay and rest states.

**Fig. 2.**
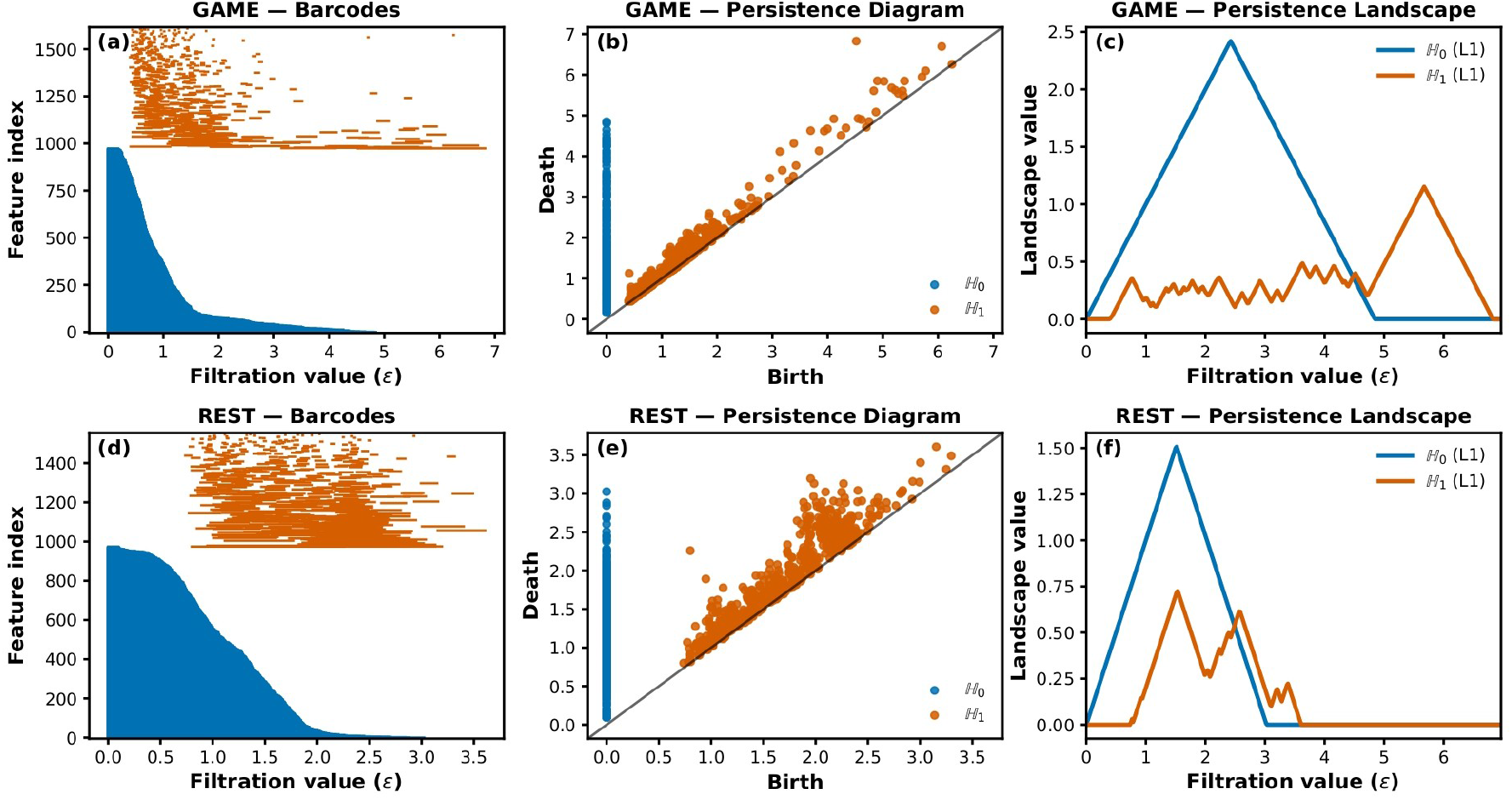
Topological representations of EEG dynamics from **Subject 27** (alpha band, channel AF8) during **gameplay** (top row) and **rest** (bottom row). Panels (a,d) show persistence barcodes, (b,e) persistence diagrams, and (c,f) first-layer persistence landscapes. Barcodes and landscapes are plotted as functions of the filtration value (*ε*). Persistence diagrams depict birth–death pairs for connected components (ℍ _0_, blue) and loops (ℍ _1_, orange), with the diagonal indicating equal birth and death.

### D. Class Balancing and Feature Integration

To mitigate the severe class imbalance between rest and gameplay epochs, two class-balancing strategies were implemented during training: (i) oversampling with SMOTE [26] combined with cost-sensitive learning, and (ii) within-subject random undersampling [27].

In the *oversampling configuration*, SMOTE was applied exclusively within each training fold to synthetically generate minority-class (rest) samples by interpolating between nearest neighbors in the feature space. The sampling ratio was selected to increase minority-class representation without fully equalizing class sizes. The minority class was increased to approximately 50–70% of the majority class within each training fold, with the ratio fixed *a priori* across experiments. In addition, cost-sensitive learning was employed by assigning higher misclassification penalties to rest epochs via explicit class weights during classifier training. This discouraged trivial majority-class predictions and improved sensitivity to underrepresented rest states. SMOTE was performed in the ETF feature space rather than on raw EEG signals and was restricted to training folds only. Because synthetically generated samples may not correspond to physiologically realizable EEG time series, SMOTE results are interpreted as facilitating minority-class decision boundary learning rather than as evidence of additional recorded rest-state data. To address concerns regarding synthetic sample realism, we additionally report results obtained using within-subject random undersampling, which avoids synthetic data generation; although undersampling yielded lower and more variable performance as expected, the qualitative conclusions regarding imbalance-aware behavior and relative channel effects were consistent across balancing strategies. For SMOTE-based experiments, classifiers used the default decision threshold; no threshold optimization was performed to avoid introducing an additional degree of freedom alongside synthetic oversampling and cost-sensitive learning. Threshold tuning was reserved for the undersampling regime to isolate the effect of representation and balancing.

In the undersampling configuration, gameplay epochs were randomly subsampled without replacement within each subject and within each training fold to match the number of rest epochs for that subject. This strategy preserves within-subject balance and data authenticity, albeit at the cost of reduced gameplay sample diversity and statistical power.

In both configurations, each EEG epoch was represented by a unified topological feature vector composed of persistence landscapes and entropy values derived from ℍ_0_ and ℍ_1_ diagrams across four canonical EEG bands and selected EEG channels (TP9 and TP10 in the reduced configuration, and TP9, TP10, AF7, and AF8 in the full configuration). Persistence landscapes were vectorized and concatenated across bands, channels, and homology dimensions, while persistence entropy contributed one scalar per homology dimension. The final feature representation is defined as:

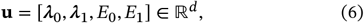

where *λ*_*k*_, *k* ∈ {0, 1}, denotes the vectorized persistence landscape functions and *E*_*k*_, *k* ∈ {0, 1}, denotes the corresponding persistence entropy. Here, *k* = 0 and *k* = 1 correspond to connected components (ℍ_0_) and one-dimensional cycles (ℍ_1_), respectively.

### E. Classification and Evaluation

Four supervised classifiers were selected to evaluate the proposed topological features across a spectrum of model complexities and interpretability. XGBoost (XGB) was chosen due to its strong performance in imbalanced, high-dimensional data settings and proven utility in EEG classification [28]. Random Forest (RF) offers resilience to noise and feature redundancy, making it well suited for topological EEG features [15]. Support Vector Machine (SVM) with an RBF kernel enables nonlinear decision boundaries, while Decision Tree (DT) provides an interpretable baseline with clear rule-based structure [29], [30].

To assess classification performance, five-fold cross-validation was conducted using the GroupKFold strategy which ensures subject-disjoint evaluation by assigning all EEG segments from a given participant to the same fold. For each fold, class balancing was applied exclusively to the training split using one of two strategies: (i) SMOTE, or (ii) random undersampling of the majority class (gameplay epochs). Test folds were left untouched. All preprocessing and feature-extraction settings were fixed *a priori* and held constant across folds. For undersampling experiments, a classifier-specific decision threshold *θ*_*k*_was optimized on the training fold to maximize balanced accuracy and then applied unchanged to the corresponding test fold. Decision threshold optimization was performed exclusively within the training fold, and no information from the test fold was used during threshold selection. Decision threshold optimization was not performed for SMOTE-based experiments to avoid compounding degrees of freedom when combining synthetic oversampling with class weighting. All preprocessing, embedding parameter estimation, feature extraction, class balancing, and decision threshold selection were performed using training data only, with no information from the test folds used at any stage.

In the SMOTE-based configuration, class weights were specified in each classifier to penalize misclassification of the minority class. Each EEG segment was represented by a feature vector of 496 or 992 dimensions, corresponding to the 2-channel and 4-channel configurations, respectively.

Model performance was quantified using accuracy, balanced accuracy, precision, recall, and F1 score. Metrics were computed independently for each class, with particular emphasis on balanced accuracy, macro-F1, and minority-class (rest) recall, which are more informative under severe class imbalance. All metrics were reported separately for training and test sets, and mean values were computed across the five cross-validation folds to ensure reliable performance estimates. All experiments were repeated under two spatial configurations:

1. A reduced 2-channel setting using TP9 and TP10, and
2. A full 4-channel setting using all Muse electrodes (TP9, TP10, AF7, AF8).

### F. Sensitivity Analysis of Topological Parameters

To assess the robustness of the proposed framework to topological feature parameterization, a linear mixed-effects (LME) analysis was conducted under the SMOTE-based training regime. The analysis evaluated the influence of persistence landscape resolution (PL) and the number of retained landscape layers (*K*) on test balanced accuracy, while accounting for repeated measurements across subject-wise cross-validation folds by treating fold as a random intercept.

Persistence landscape resolution (PL) and truncation depth (*K*) were treated as robustness factors rather than tunable hyperparameters. This analysis was designed to evaluate sensitivity to parameterization and to inform the selection of fixed values for subsequent experiments.

### G. Implementation Details, Computational Complexity, and Runtime

EEG recordings were segmented into non-overlapping 4.5 s epochs (990 samples at 220 Hz). Within each epoch, sliding windows of 330 samples with a step size of 110 samples were extracted, yielding seven windows per epoch. Phase-space reconstruction (PSR) was performed using fixed parameters (*m, τ*) = (4, 6), producing embedded point clouds of 315 points in ℝ ^4^ per window. Persistent homology was computed up to dimension *k* = 1 for each window, frequency band, and channel. Persistence landscapes and entropy values derived from ℍ_0_ and ℍ_1_ were then averaged across windows to obtain a single feature vector per epoch, resulting in feature dimensionalities of 496 and 992 for the two- and four-channel configurations, respectively.

The dominant computational cost arises from persistent homology computation on the embedded point clouds. For each retained epoch, persistence was computed for each band, sliding window, and channel (4 bands × 7 windows × 2 channels = 56 computations in the two-channel setting; 4 bands × 7 windows × 4 channels = 112 computations in the four-channel setting). Each computation returned diagrams up to dimension one (ℍ _0_ and ℍ _1_). Although Vietoris–Rips persistence has exponential worst-case complexity with respect to the number of points, practical runtime remained tractable for point clouds of size 315 using optimized implementations.

All feature extraction and classification procedures were implemented in Python 3.12 using standard scientific computing libraries, including NumPy, SciPy, scikit-learn, and matplotlib. Topological features were computed using `ripser.py` and `giotto-tda`, while data balancing was handled using `imblearn`. Experiments were executed on a workstation equipped with an Intel Core i7-11800H CPU (8 cores, 2.30 GHz) and 32 GB DDR4 RAM.

Empirical runtime was evaluated by timing end-to-end feature extraction on randomly selected EEG epochs using the same configuration employed in the main experiments (four channels, PL = 30, *K* = 1), with parallel execution across available CPU cores. Averaged over 200 epochs, the ETF pipeline required 676 ms per 4.5 s EEG epoch (≈6.7× faster than real time). While evaluated in an offline per-epoch setting, these results suggest that ETF is computationally efficient and compatible with online or near–real-time mobile EEG analysis without GPU acceleration.

## IV. Results

### A. Sensitivity Analysis of Topological Parameters

Prior to reporting classification performance, sensitivity of the proposed Enriched Topological Feature (ETF) representation to persistence landscape resolution (PL) and truncation depth (*K*) was evaluated using linear mixed-effects (LME) models on fold-wise test metrics under SMOTE-based training. Across both two-channel and four-channel configurations, no significant main effect of PL was observed for test balanced accuracy, macro-F1, or overall accuracy, nor was any PL×*K* interaction detected (*p* > 0.1 in all cases). Performance variation across the evaluated PL range (10 to 60) remained limited (mean spreads < 4 percentage points). In contrast, *K* exhibited a modest but statistically significant effect on imbalance-aware metrics, driven primarily by performance degradation at higher-order landscapes rather than consistent improvement. No monotonic gains were observed beyond *K* = 1. Accordingly, PL was fixed to 30 and *K* to 1 for all subsequent experiments, providing sufficient numerical resolution with negligible additional computational overhead.

### B. Imbalance-Aware Evaluation of ETF

Given the severe class imbalance (rest:game ≈ 1:9.3) and subject-disjoint evaluation, overall accuracy can be dominated by majority-class behavior. Accordingly, balanced accuracy, macro-F1, and minority-class (rest) recall are treated as primary outcomes, while accuracy-centric metrics are reported separately for contextual comparison. All imbalance-aware results are reported as mean and 95% confidence intervals (CI) across five subject-wise GroupKFold folds. Confidence intervals were computed using a two-sided *t*-interval over fold-wise metric estimates (*n* = 5 folds).

To verify that epoch-level fold metrics were not dominated by subjects contributing many epochs, we performed an additional subject-level sensitivity analysis. Specifically, out-of-fold epoch predictions were first aggregated within each test subject to compute per-subject balanced accuracy, macro-F1, and class-wise metrics, and these subject-level scores were then averaged across subjects with 95% bootstrap confidence intervals. The resulting subject-level estimates (Supplementary Table S5) preserve the same qualitative trends across classifiers and channel configurations, supporting that the reported performance is not driven by unequal epoch counts per subject.

Under subject-disjoint evaluation with severe class imbalance and unconstrained motion, these performance levels reflect recovery of task-relevant structure under conditions where chance-level balanced accuracy is 50%. Because rest constitutes only ∼ 9.3% of epochs, overall accuracy is dominated by majority-class performance. A trivial classifier that always predicts gameplay achieves ≈90.7% accuracy and ≈95.1% gameplay F1 while yielding 0% rest recall (Supplementary Table S2). Accordingly, accuracy values in the 85–92% range are achievable without recovering any rest-state sensitivity and should not be interpreted as evidence of class-balanced discrimination. In contrast, balanced accuracy and macro-F1 penalize this failure mode by weighting both classes.

1. *SMOTE-Based Training:* Under SMOTE-based training, ETF yields balanced accuracies consistently above chance across classifiers and channel configurations, with tree-based models (Random Forest and XGBoost) outperforming margin-based methods. In the four-channel setting, XGBoost achieves a test balanced accuracy of 62.34% [95% CI: 55.31 to 69.37] and macro-F1 of 63.47% [55.58 to 71.36], with rest recall of 34.88% [24.09 to 45.68]. In contrast, the two-channel configuration attains comparable or higher balanced accuracy, with Random Forest achieving 65.39% [58.39 to 72.40] and substantially higher rest recall of 49.51% [43.76 to 55.26] (Table I). The two-channel configuration performs comparably or better on imbalance-aware metrics, indicating that inclusion of AF7/AF8 does not reliably improve cross-subject minority-class generalization under these recording conditions. This may reflect increased sensitivity of frontal channels to motion-related contamination during free play, but this effect was not directly quantified in the present study. Consistent with this observation, gains in rest recall are largest in the TP9/TP10 configuration despite similar majority-class performance (Table I).
2. *Persistence-Only Ablation (PL vs ETF):* To assess whether persistence landscapes alone are sufficient under imbalance-aware evaluation, we repeated the SMOTE-based, subject-disjoint analysis using persistence landscape features only, while retaining the same windowing and temporal averaging scheme and removing persistence entropy features. While PL-only representations maintained competitive test accuracy and strong majority-class performance, they consistently degraded class-balanced discrimination across both channel configurations, as reflected by reduced macro-F1 and balanced accuracy. Although rest recall occasionally increased for specific classifiers, this occurred alongside a marked drop in rest F1, indicating increased minority predictions with low precision (i.e., more false positives), consistent with unstable minority-class decision boundaries. In contrast, ETF preserves minority-class sensitivity while maintaining balanced performance, indicating that persistence landscapes alone are insufficient under severe class imbalance and that entropy augmentation supports more stable imbalance-aware discrimination (see Supplementary Table S1).
3. *Within-Subject Random Undersampling:* Relative to SMOTE, undersampling generally reduces macro-F1 and yields wider confidence intervals, consistent with the loss of majority-class diversity after truncation. While minority-class recall can increase for certain classifier and configuration combinations, this increase reflects classifier bias toward the minority class (e.g., in decision trees) at the expense of majority-class precision, resulting in reduced macro-F1 and balanced accuracy. Overall, these tradeoffs support SMOTE as the more stable strategy for this dataset (see Table II).

**TABLE I.** ETF performance under SMOTE-based training (PL = 30, *K* = 1)

| Ch. | C. | Test BAcc | Test Macro-F1 | Rest Rec | Rest F1 | Game F1 |
| --- | --- | --- | --- | --- | --- | --- |
| A4 | RF | 57.39 [45.98, 68.81] | 57.97 [45.80, 70.15] | 32.32 [20.74, 43.90] | 33.99 [16.79, 51.20] | 81.95 [74.22, 89.68] |
|  | XGB | 62.34 [55.31, 69.37] | **63.47** [55.58, 71.36] | 34.88 [24.09, 45.68] | 40.37 [26.99, 53.74] | **86.58** [83.72, 89.44] |
|  | SVM | **63.53** [52.48, 74.58] | 61.98 [47.82, 76.14] | **44.17** [26.89, 61.44] | **42.09** [15.41, 68.77] | 79.80 [68.16, 91.43] |
|  | DT | 55.97 [51.04, 60.90] | 53.52 [48.79, 58.24] | 33.79 [25.66, 41.93] | 26.99 [19.33, 34.65] | 73.24 [71.03, 75.45] |
| T2 | RF | 65.39 [58.39, 72.40] | **64.48** [56.09, 72.86] | **49.51** [43.76, 55.26] | **45.71** [34.25, 57.18] | 83.24 [77.32, 89.17] |
|  | XGB | 61.68 [57.54, 65.82] | 62.01 [56.46, 67.56] | 39.97 [32.76, 47.19] | 38.61 [34.51, 42.71] | **84.05** [78.87, 89.23] |
|  | SVM | **67.53** [57.46, 77.61] | 64.20 [53.81, 74.59] | 48.19 [34.03, 62.35] | 40.24 [26.06, 54.42] | 80.21 [71.98, 88.43] |
|  | DT | 57.28 [50.26, 64.29] | 53.94 [47.39, 60.48] | 35.68 [25.76, 45.61] | 27.81 [19.13, 36.48] | 72.19 [67.90, 76.48] |

Values are mean [95% confidence interval] across five subject-wise folds. BAcc denotes balanced accuracy. Ch. denotes EEG channel configuration (A4: TP9, TP10, AF7, AF8; T2: TP9, TP10). C. denotes classifier. Rest and Game metrics correspond to the minority and majority classes, respectively (rest is the minority class). Confidence intervals were computed using a two-sided $t$ -interval over fold-wise metric estimates ( $n = 5$ ).

**TABLE II.** ETF performance under within-subject random undersampling applied to the training fold only (PL = 30, *K* = 1).

| Ch. | C. | Test BAcc | Test Macro-F1 | Rest Rec | Rest F1 | Game F1 |
| --- | --- | --- | --- | --- | --- | --- |
| A4 | RF | 58.58 [43.54, 73.61] | 58.10 [42.27, 73.93] | 41.32 [25.18, 57.47] | 37.87 [18.94, 56.80] | 78.33 [64.51, 92.15] |
|  | XGB | 60.04 [53.51, 66.57] | 60.77 [53.36, 68.18] | 31.81 [19.40, 44.21] | 36.25 [23.52, 48.97] | 85.30 [80.90, 89.70] |
|  | SVM | 65.72 [54.47, 76.96] | 59.29 [48.21, 70.36] | 45.08 [29.96, 60.20] | 34.08 [20.13, 48.04] | 73.49 [65.95, 81.03] |
|  | DT | 57.89 [51.07, 64.71] | 52.60 [45.16, 60.03] | 58.15 [47.43, 68.87] | 27.53 [18.46, 36.60] | 68.08 [61.78, 74.39] |
| T2 | RF | 62.71 [54.79, 70.63] | 62.24 [51.92, 72.57] | 44.36 [33.84, 54.87] | 42.22 [28.74, 55.70] | 82.26 [73.18, 91.34] |
|  | XGB | 60.82 [56.97, 64.67] | 61.52 [56.80, 66.24] | 33.09 [22.21, 43.97] | 37.49 [29.18, 45.80] | 85.56 [81.17, 89.95] |
|  | SVM | 64.54 [52.91, 76.17] | 60.92 [48.40, 73.43] | 59.00 [43.76, 74.24] | 37.06 [21.45, 52.68] | 77.04 [66.22, 87.87] |
|  | DT | 57.94 [53.01, 62.88] | 52.37 [46.89, 57.85] | 58.43 [51.16, 65.70] | 27.29 [22.13, 32.44] | 67.82 [60.33, 75.30] |

Values are mean [95% confidence interval] across five subject-wise folds. BAcc denotes balanced accuracy. Ch. denotes EEG channel configuration (A4: TP9, TP10, AF7, AF8; T2: TP9, TP10). C. denotes classifier. Rest and Game metrics correspond to the minority and majority classes, respectively (rest is the minority class). Confidence intervals were computed using a two-sided $t$ -interval over fold-wise metric estimates ( $n = 5$ ).

### C. Interpretation Relative to Prior Accuracy-Centric Reporting

Prior work on pediatric mobile EEG gameplay–rest classification has predominantly reported overall accuracy and majority-class (game) precision, recall, and F1 score, without evaluating minority-class (rest) sensitivity or reporting imbalance-aware metrics. Under severe class imbalance, such accuracy-centric reporting can substantially overestimate discrimination performance, as high accuracy is achievable by trivial majority-class prediction.

Accordingly, lower overall accuracy observed under the present imbalance-aware, subject-disjoint evaluation should not be interpreted as inferior performance, but rather as a direct consequence of penalizing majority-class dominance to recover minority-class sensitivity. In contrast to accuracy-centric evaluation, ETF is assessed using balanced accuracy, macro-F1, and rest recall (Tables I–II), which provide a more informative measure of class-balanced discrimination under severe imbalance.

Notably, ETF recovers rest-state sensitivity using as few as two channels (TP9/TP10), indicating that task-relevant topological structure remains informative even under low-density mobile EEG constraints. For protocol comparability only, Table S2 reports performance using the same accuracy-centric metrics adopted in prior work. Because these metrics do not evaluate minority-class sensitivity, Supplementary Table S2 should not be interpreted as an imbalance-aware benchmark and is included solely to facilitate direct comparison with previously published results.

## VII. Feature Analysis

These attribution patterns are model-dependent and should be interpreted as feature-level correlates rather than evidence of underlying oscillatory generators or causal neural mechanisms. To interpret the contribution of individual topological features to model decisions, SHAP (SHapley Additive exPlanations) analysis [31] was performed using TreeExplainer on the trained XGBoost classifier. Features were derived from the ETF pipeline applied to each EEG epoch. SHAP values quantify feature contributions to the classifier’s decision function.

Mean absolute SHAP values were used to rank feature importance across the dataset. For interpretability, features were labeled according to their frequency band, electrode location, homology dimension, and feature type (persistence landscape component or entropy).

### A. Key Topological Features

Fig. 3(a) and 3(b) summarize the most influential features identified by SHAP analysis. Overall, gameplay classification was primarily attributed by the model to gamma-band persistence landscape features at AF7/AF8 electrodes, whereas resting-state predictions were associated with alpha-band topological structure at TP9/TP10.

**Fig. 3.**
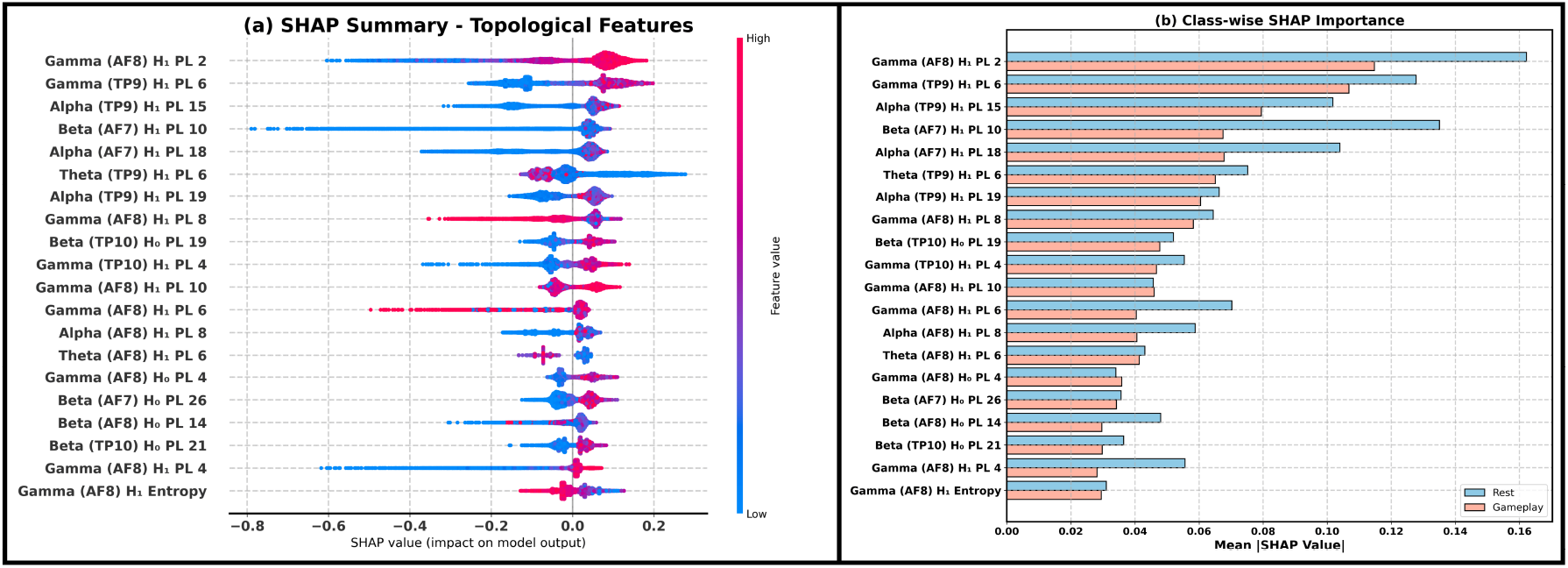
Model interpretability via SHAP-based analysis of topological EEG features. (a) SHAP summary plot showing the top 20 features ranked by impact on classifier output. Each point represents a SHAP value for an instance, colored by the corresponding feature magnitude. (b) Class-wise SHAP importance plot showing mean absolute SHAP values for rest and gameplay conditions separately. Feature labels follow the format: *Band (Channel) Homology Feature Index*. Persistence landscape indices (e.g., PL 2, PL 10) denote coordinates of the discretized persistence landscape vector (PL = 30 uniformly sampled points) and do not correspond to landscape layers or ranked topological features.

- **Gamma (AF8) ℍ** _1_ **PL 2** exhibited the highest mean absolute SHAP value (≈0.16), indicating a strong association with gameplay classification. This feature captures persistent loop-like structure in gamma-band phase-space dynamics at AF8.
- **Alpha (TP9) ℍ** _1_ **PL 15 and PL 19**, along with **Gamma (TP9) ℍ** _1_ **PL 6**, showed large negative SHAP values (mean range: –0.4 to –0.6), indicating stronger association with resting-state epochs characterized by sustained low-frequency topological organization.
- **Entropy-based features** displayed mixed SHAP polarity across bands and channels, suggesting that changes in signal complexity contributed to both gameplay and rest predictions in a state-dependent manner rather than exhibiting unidirectional effects.

### B. Neurocognitive Interpretation

The prominence of frontal gamma-band persistence landscapes, particularly at AF8, is consistent with patterns commonly reported during increased cognitive engagement in goal-directed tasks. Gamma activity has been associated in prior work with rapid cortical processing, working memory maintenance, and executive control mechanisms that are heavily recruited during visually complex and goal-directed tasks such as *Minecraft* [32], [33]. Importantly, the ETF framework captures not gamma power per se, but the persistence of loop-like structures in the reconstructed phase space, reflecting stable dynamical patterns rather than transient amplitude fluctuations.

In contrast, alpha-band persistence landscapes at TP9 were strongly linked to resting-state predictions. Alpha rhythms are known to index sensory inhibition and internally oriented processing, and their sustained topological structure is consistent with stable default-mode network dynamics during cognitive disengagement [34].

Lateralized effects were also observed in the beta band. **Beta (AF7) ℍ** _1_ **PL 10** contributed positively to gameplay classification, whereas homologous right-sided TP10 features showed weaker influence. This asymmetry is compatible with left-hemisphere dominance in visuomotor planning and top-down control, and aligns with prior evidence of beta–gamma interactions during motor coordination and task execution [35].

### C. Topological Signatures of Cognitive Load

Beyond individual features, SHAP analysis revealed systematic differences in topological organization between behavioral states. Reduced alpha-band entropy during gameplay suggests a relative suppression of spontaneous cortical variability, consistent with constrained neural dynamics under elevated executive demand [36]. Concurrently, increased persistence of gamma-band ℍ _1_ landscapes at AF7/AF8 electrodes reflects more stable task-associated dynamical organization in the reconstructed phase space.

While high frequency EEG components in mobile settings may be susceptible to myogenic contamination, the dominance of persistence based features rather than raw spectral power suggests that classification relies on the stability of geometric structure in phase space. Nonetheless, gamma related findings are interpreted conservatively as markers of task associated dynamical organization rather than exclusively neurogenic oscillatory activity.

### D. Clinical and Developmental Relevance

The observed gamma–alpha dissociation and hemispheric asymmetries are particularly relevant in the context of pediatric neurodevelopment [37]. Frontal gamma persistence during gameplay may reflect compensatory or immature control strategies in developing cortico-cortical networks engaged during top-down regulation. Beta-band lateralization further implicates frontoparietal and premotor pathways involved in visuomotor integration and early-stage motor learning. Together, these results indicate that ETF captures dynamical features that vary with developmental factors and are not fully described by conventional spectral descriptors. These findings are intended to support clinician-facing monitoring and decision support in naturalistic settings rather than diagnostic classification or clinical state labeling.

These attributions describe model reliance on engineered topological descriptors, not source-localized generators or causal neural mechanisms.

## VIII. Discussion

### A. Implications Under Deployment Constraints

This study introduces *Enriched Topological Features* (ETF) as a feature-level interpretable framework for classifying cognitive brain states from low-density pediatric EEG acquired in naturalistic settings. By integrating persistent homology, phase-space embeddings, and time-aggregated entropy, ETF supports reliable gameplay–rest discrimination under subject-disjoint evaluation, severe class imbalance, and limited spatial resolution.

Rather than optimizing accuracy alone, ETF emphasizes imbalance-aware performance, demonstrating stable class-balanced discrimination and recovery of minority-class (rest) sensitivity while maintaining robust majority-class performance across classifiers and channel configurations. These findings indicate that topological representations can remain informative under motion, environmental noise, and participant movement, supporting practical use in mobile pediatric EEG settings.

Under these deployment-relevant constraints, performance remains moderate, with best test balanced accuracy in the range of approximately 65 to 67% and minority-class (rest) recall reaching up to approximately 50% in the TP9/TP10 configuration. These values are expected given the combination of unconstrained movement, low channel count, severe class imbalance, and subject heterogeneity enforced by subject-disjoint evaluation. The practical implication is support for coarse engagement–rest monitoring rather than fine-grained cognitive state decoding, with recovery of minority-class sensitivity constituting the primary limiting factor for real-world deployment.

### B. Interpretability and Neurophysiological Context

Model attribution analysis using SHAP revealed structured topological patterns that differentiate gameplay engagement from rest. Gameplay epochs were consistently associated with increased contributions from gamma-band persistence landscape features at AF7/AF8 electrodes, most prominently AF8 in ℍ _1_. These features are consistent with enhanced dynamical organization during task engagement, consistent with prior associations between gamma-band activity and fast cortical processing, working memory, and executive control during goal-directed behavior [38]. Importantly, these observations represent model attribution patterns rather than direct neurobiological measurements.

In contrast, resting-state predictions were dominated by alpha-band entropy and persistence features at TP9. This pattern is consistent with metastable disengagement and internally oriented processing, aligning with established roles of alpha rhythms in sensory inhibition and default-mode network dynamics [39].

Lateralized asymmetries were also observed across beta and gamma features, with stronger task-related contributions arising from left frontal regions compared to right-sided TP10 features. Such asymmetries are consistent with developmental neurophysiology, wherein frontoparietal and parietal–cerebellar pathways support visuomotor coordination and executive–motor integration during childhood [40]. Collectively, these attribution patterns suggest that ETF captures state-dependent reconfigurations of neural dynamics rather than incidental signal fluctuations or classifier artifacts.

### C. Developmental Trends and Exploratory Analysis

This exploratory analysis used SHAP-guided feature selection and was not corrected for multiple comparisons. To examine potential developmental associations, we analyzed the relationship between participant age and gamma-band topological features during gameplay. For each subject (*N* = 43), features were averaged across all retained gameplay epochs to obtain a single subject-level estimate. The analysis focused on the mean lifetime of the two most persistent ℍ _1_ loops in the frontal gamma band (30 to 35 Hz, AF8), a persistence-landscape–derived measure selected based on SHAP analysis as the most influential gamma-band feature for gameplay classification.

At the subject level, no statistically significant association was observed between age and frontal gamma-band ℍ _1_ loop persistence (Pearson *r* = 0.05, *p* = 0.77; Spearman *ρ* = 0.11, *p* = 0.50). Bootstrap resampling yielded wide confidence intervals spanning zero (Pearson 95% CI: [− 0.20, 0.36]), indicating substantial inter-individual variability.

From a neurodevelopmental perspective, models of synaptic pruning and large-scale network maturation predict a transition from locally recurrent to more efficient network organization across development [37], [41]. Such models would be expected to reduce the persistence of high-frequency reverberatory dynamics with age [42]. However, no statistically detectable age-related modulation of the extracted gamma-band topological feature was observed in the present cohort, suggesting relative developmental stability under naturalistic, low-density mobile EEG recording conditions.

To assess potential motion-related confounding, we performed an accelerometer-based control analysis. Within-subject correlations between epoch-level gamma-band ℍ _1_ loop persistence and the root-mean-square of the DC-removed triaxial accelerometer magnitude were centered near zero (mean *r* = 0.007, bootstrap 95% CI: [− 0.06, 0.07]). Controlling for mean gameplay motion at the subject level did not materially alter the age–feature relationship (*r* = 0.09, *p* = 0.57). Finally, no significant differences in frontal gamma-band ℍ _1_ loop persistence were observed across self-reported Minecraft gameplay frequency groups (Never, Sometimes, Often; one-way ANOVA, *F* = 0.20, *p* = 0.82), indicating no detectable modulation by gameplay exposure within this cohort.

### D. Relation to EEG Microstate Dynamics

EEG microstate analysis models scalp activity as a sequence of short-lived, quasi-stable spatial configurations and is commonly used to summarize large-scale temporal organization in cognitive and developmental EEG [43]. Prior work reports that increased attentional demand is often accompanied by shifts in microstate expression and reduced dominance of default-mode–like patterns, whereas resting conditions show longer state lifetimes and greater default-mode occupancy [44].

We do not claim microstate validation in this work; the following comparison is interpretive only. Because microstates cannot be reliably estimated from the present four-channel montage, the microstate literature is used solely as a conceptual reference for interpreting state-dependent differences in temporal organization. Consistent with the general contrast between task-driven and internally oriented dynamics, gameplay epochs were characterized by increased frontal gamma-band ℍ _1_ persistence and reduced alpha-band structure at TP9/TP10, whereas rest epochs showed stronger alpha-band topological organization. These patterns qualitatively align with reports of task-related reductions in default-mode–like dynamics in microstate studies.

Overall, microstate and topological analyses emphasize complementary levels of description. Microstates summarize recurrent scalp-level spatial configurations when sufficient spatial sampling is available, whereas ETF characterizes the geometric organization of band-limited single-channel dynamics in a manner that remains informative under low-density mobile EEG.

### E. Clinical Implications and Real-World Applicability

ETF is motivated by real-world pediatric settings where traditional high-density EEG is impractical due to cost, setup complexity, and poor tolerance to movement. Potential applications include monitoring cognitive engagement during serious gaming or neurorehabilitation and supporting adaptive task design under ecologically valid conditions. Importantly, ETF is intended as a decision-support and monitoring framework rather than a diagnostic tool, enabling coarse but robust differentiation of cognitive states from low-density mobile EEG.

From a computational standpoint, ETF demonstrates favorable efficiency for resource-constrained settings. End-to-end feature extraction required an average of 676 ms per 4.5 s EEG epoch using four channels (PL = 30, *K* = 1) with parallel CPU execution (≈6.7× faster than real time). Although runtime was evaluated offline on a per-epoch basis, these results support near–realtime feasibility without GPU acceleration. These measurements reflect full per-epoch processing, including filtering, phase-space reconstruction, persistent homology, and feature aggregation, and support near–real-time operation on standard multi-core hardware.

From a translational perspective, ETF is best viewed as a clinician- or therapist-facing monitoring tool rather than an autonomous decision system. In practical scenarios, ETF-derived metrics could be computed continuously during neurorehabilitation or serious gaming sessions to quantify engagement-related neural organization. Such measures may inform adaptive task difficulty or session pacing, or support early identification of disengagement, particularly in pediatric populations where sustained attention is variable.

### F. Methodological Limitations

Several limitations should be acknowledged. First, the use of a four-channel EEG system constrains spatial resolution, limiting access to distributed cortical networks and precluding source-level analysis. While this design choice enhances ecological validity and scalability in pediatric settings, ETF should be viewed as complementary to high-density EEG rather than a replacement. Second, although nested cross-validation or a held-out subject test set could be employed for strict hyperparameter optimization, the present study treats persistence landscape resolution and truncation depth as robustness parameters rather than tunable hyperparameters. The absence of performance sensitivity across the evaluated parameter range supports the stability of the proposed representation and reduces the risk of configuration-specific overfitting.

Third, the binary classification paradigm adopted here does not capture finer-grained cognitive states such as flow, frustration, or partial disengagement during gameplay. Extension to multi-class or ordinal state classification represents an important direction for future work.

Finally, interpretation of high-frequency EEG features in mobile pediatric contexts warrants caution. Although accelerometer-based analyses using the root-mean-square of the DC-removed triaxial motion magnitude revealed no systematic linear association between head motion and gamma-band topological features across participants (mean within-subject *r* = 0.007, bootstrap 95% CI: [− 0.06, 0.07]), gamma-band activity remains susceptible to subtle myogenic contamination [45]. While reliance on persistence-based topological features may provide partial robustness to transient artifacts, definitive separation of neurogenic and myogenic sources will require concurrent EMG monitoring or multimodal artifact characterization. Although quality-control procedures and the use of persistence-based descriptors may reduce sensitivity to transient artifacts, high-frequency EEG components in mobile pediatric recordings remain susceptible to myogenic contamination. Therefore, gamma-band findings are interpreted conservatively as gamma-band–filtered topological dynamics rather than direct evidence of neurogenic gamma oscillations.

### G. Future Directions

Future work should prioritize large-scale validation studies with target sample sizes of approximately 150–250 participants to assess generalizability and to establish age-adjusted normative ranges for ETF-derived features. Multi-center collaborations employing harmonized MoBI-style acquisition protocols will be critical for evaluating robustness across recording environments, hardware platforms, and population heterogeneity. Methodologically, extensions to ordinal or multi-class classification frameworks may enable characterization of intermediate cognitive states beyond binary rest and gameplay conditions, such as graded engagement or partial disengagement. Such extensions would provide a more nuanced representation of cognitive dynamics in naturalistic settings. Finally, integrating ETF with complementary EEG analysis paradigms, including microstate analysis or connectivity-based representations, may support a multilevel description of pediatric brain dynamics. Linking transient scalp-level patterns to underlying phase-space topology could offer deeper insight into the temporal organization of neural activity during development.

## IX. Conclusion

This study establishes a deployment-relevant baseline for pediatric mobile EEG gameplay–rest classification by reporting what performance remains under subject-disjoint evaluation with untouched, naturally imbalanced test folds. Under these constraints, Enriched Topological Features (ETF) enable stable class-balanced discrimination and recovery of minority-class (rest) sensitivity with near–real-time computational feasibility.

ETF provides a computationally efficient and interpretable framework for classifying pediatric EEG recorded in naturalistic settings. By integrating persistent homology, time-aggregated entropy, and phase-space embeddings, ETF captures structured, state-dependent neural dynamics that remain informative under low spatial resolution, substantial motion, and severe class imbalance, without relying on accuracy-centric evaluation.

Taken together, the proposed framework supports pediatric cognitive-state monitoring and longitudinal developmental research using low-density mobile EEG, with potential utility in real-world educational and neurorehabilitation contexts.

## Data Availability

All data produced in the present study are available upon reasonable request to the authors

## References

[1] E. S. Norton et al., “Social EEG: A novel neurodevelopmental approach to studying brain-behavior links and brain-to-brain synchrony during naturalistic toddler–parent interactions,” Dev. Psychobiol., vol. 64, no. 3, p. e22240, 2022.

[2] A. Giannadou et al., “Investigating neural dynamics in autism spectrum conditions outside of the laboratory using mobile electroencephalography,” Psychophysiology, vol. 59, no. 4, p. e13995, 2022.

[3] S. Georgieva et al., “Toward the understanding of topographical and spectral signatures of infant movement artifacts in naturalistic EEG,” Front. Neurosci., vol. 14, p. 352, 2020.

[4] S. Makeig et al., “Linking brain, mind and behavior,” Proc. IEEE, vol. 100, no. Special Centennial Issue, pp. 528–541, 2009.

[5] K. Gramann et al., “Cognition in action: Imaging brain/body dynamics in mobile humans,” Rev. Neurosci., vol. 25, no. 3, pp. 365–380, 2014.

[6] S. Ladouce et al., “Understanding minds in real-world environments: Toward a mobile cognition approach,” Front. Hum. Neurosci., vol. 10, p. 694, 2017.

[7] A. S. Ravindran et al., “Assaying neural activity of children during video game play in public spaces: A deep learning approach,” J. Neural Eng., vol. 16, no. 3, p. 036028, 2019.

[8] Z. Raeisi et al., “EEG microstate biomarkers for schizophrenia: A novel approach using deep neural networks,” Cogn. Neurodyn., vol. 19, no. 1, pp. 1–26, 2025.

[9] Z. Raeisi et al., “Enhanced classification of tinnitus patients using EEG microstates and deep learning techniques,” Sci. Rep., vol. 15, no. 1, p. 15959, 2025.

[10] R. A. Lashaki et al., “EEG microstate analysis in trigeminal neuralgia: Identifying potential biomarkers for enhanced diagnostic accuracy,” Acta Neurol. Belg., pp. 1–21, 2025.

[11] M. Rahmati et al., “Novel EEG-based diagnostic framework for major depressive disorder using microstate and entropy features,” Cogn. Neurodyn., vol. 19, no. 1, p. 116, 2025.

[12] M. Saggar et al., “Towards a new approach to reveal dynamical organization of the brain using topological data analysis,” Nat. Commun., vol. 9, no. 1, p. 1399, 2018.

[13] X. Xu et al., “Topological data analysis as a new tool for EEG processing,” Front. Neurosci., vol. 15, p. 761703, 2021.

[14] N. Aithal et al., “Leveraging persistent homology for differential diagnosis of mild cognitive impairment,” in Proc. Int. Conf. Pattern Recognit., 2025, pp. 17–32.

[15] K. Gaurav et al., “Characterizing neural activity during video game engagement using EEG sensor based topological dynamics analysis,” IEEE Sens. Lett., 2024.

[16] A. Zomorodian and G. Carlsson, “Computing persistent homology,” in Proc. 20th Annu. Symp. Comput. Geom., 2004, pp. 347–356.

[17] H. Edelsbrunner et al., “Topological persistence and simplification,” Discrete Comput. Geom., vol. 28, pp. 511–533, 2002.

[18] P. Bubenik et al., “Statistical topological data analysis using persistence landscapes,” J. Mach. Learn. Res., vol. 16, no. 1, pp. 77–102, 2015.

[19] N. Atienza et al., “Persistent entropy for separating topological features from noise in Vietoris-Rips complexes,” J. Intell. Inf. Syst., vol. 52, pp. 637–655, 2019.

[20] A. S. Ravindran et al., “Multi-modal mobile brain-body imaging (MoBI) dataset for assaying neural and head movement responses associated with creative video game play in children,” IEEE Dataport, 2017, [Online]. Available: doi: 10.21227/H23W88.

[21] A. Widmann et al., “Digital filter design for electrophysiological data: A practical approach,” J. Neurosci. Methods, vol. 250, pp. 34–46, 2015.

[22] P. Welch, “The use of fast Fourier transform for the estimation of power spectra: A method based on time averaging over short, modified periodograms,” IEEE Trans. Audio Electroacoust., vol. 15, no. 2, pp. 70–73, 1967.

[23] F. Takens, “Detecting strange attractors in turbulence,” in Dynamical Systems and Turbulence, Warwick 1980, 2006, pp. 366–381.

[24] A. M. Fraser and H. L. Swinney, “Independent coordinates for strange attractors from mutual information,” Phys. Rev. A, vol. 33, no. 2, pp. 1134–1140, 1986.

[25] M. B. Kennel et al., “Determining embedding dimension for phasespace reconstruction using a geometrical construction,” Phys. Rev. A, vol. 45, no. 6, pp. 3403–3411, 1992.

[26] N. V. Chawla et al., “SMOTE: Synthetic minority over-sampling technique,” J. Artif. Intell. Res., vol. 16, pp. 321–357, 2002.

[27] H. He and E. A. Garcia, “Learning from imbalanced data,” IEEE Trans. Knowl. Data Eng., vol. 21, no. 9, pp. 1263–1284, 2009.

[28] A. Shen et al., “A data-centric and interpretable EEG framework for depression severity grading using SHAP-based insights,” J. Neuroeng. Rehabil., vol. 22, no. 1, pp. 1–16, 2025.

[29] F. Lotte et al., “A review of classification algorithms for EEG-based brain–computer interfaces: A 10 year update,” J. Neural Eng., vol. 15, no. 3, p. 031005, 2018.

[30] L. Breiman, “Random forests,” Mach. Learn., vol. 45, pp. 5–32, 2001.

[31] S. M. Lundberg and S.-I. Lee, “A unified approach to interpreting model predictions,” Adv. Neural Inf. Process. Syst., vol. 30, 2017.

[32] M. W. Howard et al., “Gamma oscillations correlate with working memory load in humans,” Cereb. Cortex, vol. 13, no. 12, pp. 1369– 1374, 2003.

[33] F. Aoki et al., “Increased gamma-range activity in human sensorimotor cortex during performance of visuomotor tasks,” Clin. Neurophysiol., vol. 110, no. 3, pp. 524–537, 1999.

[34] L. Pei et al., “Comparative analysis of multifaceted neural effects associated with varying endogenous cognitive load,” Commun. Biol., vol. 6, no. 1, p. 795, 2023.

[35] P.-M. Matta et al., “Modulation of beta oscillatory dynamics in motor and frontal areas during physical fatigue,” Commun. Biol., vol. 8, no. 1, p. 687, 2025.

[36] W. Klimesch et al., “EEG alpha oscillations: The inhibition–timing hypothesis,” Brain Res. Rev., vol. 53, no. 1, pp. 63–88, 2007.

[37] P. J. Uhlhaas et al., “The development of neural synchrony reflects late maturation and restructuring of functional networks in humans,” Proc. Natl. Acad. Sci. U.S.A., vol. 106, no. 24, pp. 9866–9871, 2009.

[38] M. Assem et al., “High gamma activity distinguishes frontal cognitive control regions from adjacent cortical networks,” Cortex, vol. 159, pp. 286–298, 2023.

[39] Y.-W. Kim et al., “The importance of low-frequency alpha (8–10 Hz) waves and default mode network in behavioral inhibition,” Clin. Psychopharmacol. Neurosci., vol. 22, no. 1, pp. 53–62, 2023.

[40] E. Ünsal et al., “From infancy to childhood: A comprehensive review of event-and task-related brain oscillations,” Brain Sci., vol. 14, no. 8, p. 837, 2024.

[41] D. A. Fair et al., “Functional brain networks develop from a “local to distributed” organization,” PLoS Comput. Biol., vol. 5, no. 5, p. e1000381, 2009.

[42] S. D. McKeon et al., “Age-related differences in transient gamma band activity during working memory maintenance through adolescence,” Neuroimage, vol. 274, p. 120112, 2023.

[43] C. M. Michel and T. Koenig, “EEG microstates as a tool for studying the temporal dynamics of whole-brain neuronal networks: A review,” Neuroimage, vol. 180, pp. 577–593, 2018.

[44] B. A. Seitzman et al., “Cognitive manipulation of brain electric microstates,” Neuroimage, vol. 146, pp. 533–543, 2017.

[45] A. J. Shackman et al., “Electromyogenic artifacts and electroencephalographic inferences,” Brain Topogr., vol. 22, pp. 7–12, 2009.

