## Supplementary Data for "Subject-Disjoint, Imbalance-Aware Gameplay–Rest Classification in Pediatric Mobile EEG Using Enriched Topological Features"

TABLE S1: Persistence-only ablation under SMOTE-based training (PL = 30, $K = 1$). Performance is reported using imbalance-aware metrics under subject-disjoint evaluation. PL-only denotes persistence landscape features computed with the same sliding-window extraction and temporal averaging scheme as ETF, but without persistence entropy features.

| Ch. | C. | Test BAcc | Test Macro-F1 | Rest Rec | Rest F1 | Game F1 |
|---|---|---|---|---|---|---|
| A4 | RF | 53.67 [51.13, 56.20] | 51.27 [48.82, 53.71] | 26.95 [19.42, 34.49] | 17.21 [13.02, 21.41] | 85.32 [81.99, 88.65] |
| | XGB | 54.81 [51.57, 58.06] | **54.20** [52.10, 56.30] | 19.88 [12.34, 27.43] | 17.97 [13.57, 22.36] | **90.43** [88.80, 92.07] |
| | SVM | **60.65** [56.39, 64.90] | 54.16 [51.37, 56.95] | **44.56** [32.46, 56.65] | **24.49** [22.51, 26.48] | 83.82 [77.99, 89.66] |
| | DT | 55.13 [52.82, 57.44] | 49.13 [47.44, 50.81] | 39.86 [32.59, 47.14] | 18.76 [16.45, 21.06] | 79.50 [78.03, 80.96] |
| T2 | RF | 60.52 [56.26, 64.78] | 55.99 [51.47, 60.50] | 39.45 [25.39, 53.52] | 25.41 [19.33, 31.49] | 86.57 [81.08, 92.05] |
| | XGB | 56.96 [53.58, 60.34] | 55.67 [52.56, 58.77] | 25.95 [16.54, 35.35] | 21.56 [14.38, 28.73] | **89.78** [87.45, 92.11] |
| | SVM | **64.92** [61.56, 68.27] | **56.81** [49.49, 64.13] | **53.10** [39.06, 67.15] | **29.66** [23.67, 35.64] | 83.97 [74.70, 93.24] |
| | DT | 56.96 [55.34, 58.58] | 50.60 [48.96, 52.25] | 41.97 [36.30, 47.65] | 20.53 [16.98, 24.08] | 80.68 [78.91, 82.46] |

Values are mean [95% confidence interval] across five subject-wise folds. BAcc denotes balanced accuracy. Ch. denotes EEG channel configuration (A4: TP9, TP10, AF7, AF8; T2: TP9, TP10). C. denotes classifier. Rest and Game metrics correspond to the minority (rest) and majority (game) classes, respectively. Confidence intervals were computed using a two-sided $t$-interval over fold-wise metric estimates ($n = 5$).

TABLE S2: Accuracy-centric majority-class metrics reported in prior work (not imbalance-aware). A trivial majority-class baseline is included to illustrate the inflation of accuracy-centric metrics under severe class imbalance.

| Method | Channels | Classifier | Test Acc (%) | Prec (%) | Rec (%) | F1 (%) |
|---|---|---|---|---|---|---|
| Trivial majority-class baseline (always predict gameplay) | – | – | 90.7 | 90.7 | 100.0 | 95.1 |
| DL [1] | TP9,TP10 | – | 67.40 | 67.99 | 65.55 | 66.80 |
| | All 4 | – | 73.68 | 72.91 | 71.40 | 72.15 |
| NETDA [2] | TP9,TP10 | RF | 89.71 | 90.90 | 88.46 | 89.71 |
| | | SVM | 87.96 | 91.50 | 86.71 | 87.96 |
| | | DT | 83.08 | 84.27 | 81.33 | 83.08 |
| | All 4 | RF | 91.96 | 92.72 | 91.21 | 91.96 |
| | | SVM | 89.92 | 90.68 | 89.17 | 89.92 |
| | | DT | 88.22 | 88.98 | 87.47 | 88.22 |
| ETF (this work) | TP9,TP10 | RF | 74.48 | 85.52 | 81.28 | 83.24 |
| | | XGB | 74.95 | 83.56 | 84.75 | 84.05 |
| | | SVM | 71.61 | 87.58 | 74.20 | 80.21 |
| | | DT | 61.25 | 82.85 | 64.10 | 72.19 |
| | All 4 | RF | 71.77 | 81.84 | 82.47 | 81.95 |
| | | XGB | 78.12 | 83.66 | 89.80 | 86.58 |
| | | SVM | 70.61 | 84.97 | 76.17 | 79.80 |
| | | DT | 61.93 | 81.95 | 66.26 | 73.24 |

Only metrics reported in prior work are shown. All precision, recall, and F1 values correspond exclusively to the majority (gameplay) class. Minority-class (rest) and imbalance-aware metrics are intentionally excluded and reported separately.

TABLE S3: Subject attrition from raw dataset to final paired cohort. Counts reflect successive inclusion criteria applied prior to classification. Subject exclusion was driven exclusively by missing segmentation metadata or failure to retain any quality-controlled epochs in one or both conditions.

| **Processing Stage** | **Subjects Remaining** |
|---|---|
| EEG recording files available in local dataset | 171 |
| Recordings with baseline and gameplay segmentation markers | 88 |
| Valid baseline and gameplay marker indices | 81 |
| ≥1 QC-passed resting epoch | 46 |
| ≥1 QC-passed gameplay epoch | 56 |
| ≥1 QC-passed epoch in both conditions (paired cohort) | **43** |

Quality control (QC) included amplitude-based rejection and spectral contamination screening. Paired inclusion required at least one QC-passed epoch in both resting and gameplay conditions.

TABLE S4: Subject-level phase-space embedding parameters estimated using Average Mutual Information (AMI) and False Nearest Neighbors (FNN) analysis for the retained cohort ($N = 43$). Per-channel estimates are reported for completeness, along with subject-wise medians. Channel labels follow the Muse headset convention (TP9, AF7, AF8, TP10).

| **Sub** | $\tau_{\text{TP9}}$ | $\tau_{\text{AF7}}$ | $\tau_{\text{AF8}}$ | $\tau_{\text{TP10}}$ | $\tau_{\text{med}}$ | $m_{\text{TP9}}$ | $m_{\text{AF7}}$ | $m_{\text{AF8}}$ | $m_{\text{TP10}}$ | $m_{\text{med}}$ |
|---|---|---|---|---|---|---|---|---|---|---|
| Sub 1 | 6 | 6 | 6 | 6 | 6 | 4 | 3 | 3 | 4 | 3 |
| Sub 2 | 6 | 6 | 6 | 6 | 6 | 4 | 3 | 3 | 4 | 3 |
| Sub 3 | 6 | 6 | 6 | 6 | 6 | 4 | 3 | 4 | 4 | 4 |
| Sub 4 | 6 | 6 | 6 | 6 | 6 | 4 | 3 | 3 | 4 | 3 |
| Sub 5 | 6 | 6 | 6 | 6 | 6 | 4 | 3 | 4 | 4 | 4 |
| Sub 6 | 6 | 6 | 6 | 6 | 6 | 4 | 4 | 4 | 4 | 4 |
| Sub 7 | 6 | 6 | 6 | 6 | 6 | 4 | 4 | 4 | 4 | 4 |
| Sub 8 | 6 | 6 | 6 | 6 | 6 | 4 | 4 | 4 | 4 | 4 |
| Sub 9 | 6 | 6 | 6 | 7 | 6 | 4 | 4 | 4 | 4 | 4 |
| Sub 10 | 6 | 6 | 6 | 6 | 6 | 4 | 4 | 4 | 4 | 4 |
| Sub 11 | 6 | 6 | 6 | 6 | 6 | 4 | 4 | 4 | 4 | 4 |
| Sub 13 | 6 | 6 | 8 | 6 | 6 | 4 | 4 | 4 | 4 | 4 |
| Sub 14 | 6 | 7 | 6 | 6 | 6 | 4 | 4 | 4 | 4 | 4 |
| Sub 16 | 6 | 6 | 6 | 6 | 6 | 4 | 4 | 4 | 4 | 4 |
| Sub 18 | 6 | 6 | 8 | 6 | 6 | 4 | 4 | 4 | 4 | 4 |
| Sub 21 | 6 | 7 | 6 | 6 | 6 | 4 | 4 | 4 | 4 | 4 |
| Sub 22 | 6 | 6 | 8 | 6 | 6 | 4 | 4 | 4 | 4 | 4 |
| Sub 23 | 6 | 8 | 6 | 6 | 6 | 4 | 4 | 4 | 4 | 4 |
| Sub 24 | 6 | 6 | 6 | 6 | 6 | 4 | 4 | 4 | 4 | 4 |
| Sub 26 | 6 | 6 | 6 | 6 | 6 | 4 | 4 | 4 | 4 | 4 |
| Sub 27 | 6 | 6 | 6 | 6 | 6 | 4 | 4 | 4 | 4 | 4 |
| Sub 28 | 6 | 6 | 6 | 6 | 6 | 4 | 4 | 4 | 4 | 4 |
| Sub 29 | 6 | 6 | 8 | 6 | 6 | 4 | 3 | 4 | 4 | 4 |
| Sub 31 | 6 | 7 | 6 | 6 | 6 | 4 | 4 | 4 | 4 | 4 |
| Sub 32 | 6 | 6 | 8 | 6 | 6 | 4 | 4 | 4 | 4 | 4 |
| Sub 34 | 6 | 6 | 7 | 6 | 6 | 4 | 4 | 4 | 4 | 4 |
| Sub 35 | 6 | 8 | 8 | 6 | 7 | 4 | 4 | 4 | 4 | 4 |
| Sub 36 | 6 | 6 | 6 | 6 | 6 | 4 | 4 | 4 | 4 | 4 |
| Sub 38 | 6 | 8 | 8 | 6 | 7 | 4 | 4 | 4 | 4 | 4 |
| Sub 40 | 7 | 8 | 8 | 6 | 7 | 4 | 4 | 4 | 4 | 4 |
| Sub 46 | 6 | 7 | 8 | 6 | 6 | 4 | 4 | 4 | 4 | 4 |
| Sub 48 | 6 | 6 | 6 | 6 | 6 | 4 | 4 | 4 | 4 | 4 |
| Sub 49 | 6 | 6 | 6 | 6 | 6 | 4 | 4 | 4 | 4 | 4 |
| Sub 58 | 6 | 6 | 6 | 6 | 6 | 4 | 4 | 4 | 4 | 4 |
| Sub 59 | 6 | 6 | 8 | 6 | 6 | 4 | 4 | 4 | 4 | 4 |
| Sub 61 | 6 | 6 | 6 | 6 | 6 | 4 | 4 | 4 | 4 | 4 |
| Sub 66 | 6 | 6 | 6 | 6 | 6 | 4 | 4 | 4 | 4 | 4 |
| Sub 68 | 6 | 6 | 6 | 6 | 6 | 4 | 4 | 4 | 4 | 4 |
| Sub 69 | 6 | 8 | 7 | 6 | 6 | 4 | 4 | 4 | 4 | 4 |
| Sub 75 | 6 | 6 | 8 | 6 | 6 | 4 | 4 | 4 | 4 | 4 |
| Sub 77 | 6 | 6 | 6 | 6 | 6 | 4 | 4 | 4 | 4 | 4 |
| Sub 84 | 6 | 8 | 8 | 6 | 7 | 4 | 4 | 4 | 4 | 4 |
| Sub 85 | 6 | 6 | 6 | 7 | 6 | 4 | 4 | 4 | 4 | 4 |

$\tau$ denotes the AMI-derived time delay (samples) defined as the first local minimum of the mutual information curve. $m$ denotes the embedding dimension selected via FNN using a 10% false-neighbor threshold. TP9/TP10 are left/right temporoparietal electrodes and AF7/AF8 are left/right anterior frontal electrodes. Cohort-level summaries used the subject-wise medians ($\tau_{\text{med}}$, $m_{\text{med}}$).

TABLE S5: Subject-level sensitivity analysis of ETF performance under SMOTE-based training (PL = 30, $K = 1$)

| Ch. | C. | Subj. BAcc | Subj. Macro-F1 | Rest Rec | Rest F1 | Game F1 |
|---|---|---|---|---|---|---|
| A4 | RF | 58.16 [53.21, 63.51] | 53.22 [47.87, 58.78] | 28.36 [19.00, 38.74] | 22.94 [14.73, 31.92] | 83.51 [77.92, 87.96] |
| | XGB | 56.19 [52.39, 60.34] | 52.99 [48.78, 57.22] | 18.87 [12.10, 26.63] | 20.00 [12.96, 27.90] | **85.98** [81.05, 89.95] |
| | SVM | **62.40** [57.54, 67.29] | **56.50** [51.52, 61.40] | **38.02** [28.36, 48.05] | **29.61** [21.89, 38.09] | 83.38 [77.74, 87.93] |
| | DT | 54.41 [50.00, 59.05] | 48.57 [44.86, 52.43] | 35.58 [27.48, 44.04] | 21.64 [15.39, 28.44] | 75.50 [70.32, 79.59] |
| T2 | RF | 62.64 [57.40, 68.07] | 57.66 [52.51, 63.18] | 38.68 [28.58, 48.66] | 32.01 [23.10, 41.61] | 83.32 [78.09, 87.71] |
| | XGB | 58.20 [54.74, 61.95] | 54.73 [51.11, 58.50] | 26.17 [19.36, 33.39] | 24.63 [18.09, 31.95] | **84.83** [79.63, 89.15] |
| | SVM | **64.98** [59.50, 70.27] | **57.15** [52.08, 62.67] | **47.96** [37.97, 57.56] | **33.13** [24.79, 42.34] | 81.16 [75.54, 85.89] |
| | DT | 58.14 [53.63, 63.15] | 50.21 [46.64, 54.20] | 43.33 [35.02, 52.35] | 25.10 [19.00, 31.85] | 75.31 [70.59, 79.51] |

Values are mean [95% confidence interval] computed at the subject level. Subject-level metrics were obtained by aggregating out-of-fold epoch predictions per subject, followed by bootstrap resampling over subjects. BAcc denotes balanced accuracy.

### References


[1] A. S. Ravindran, A. Mobiny, J. G. Cruz-Garza, A. Paek, A. Kopteva, and J. L. Contreras-Vidal, "Assaying neural activity of children during video game play in public spaces: A deep learning approach," *J. Neural Eng.*, vol. 16, no. 3, p. 036028, 2019.

[2] K. Gaurav, J. Landge, and T. K. R. Bollu, "Characterizing neural activity during video game engagement using EEG sensor based topological dynamics analysis," *IEEE Sens. Lett.*, 2024.
